# Adolescents’ health literacy, health protective measures, and health-related quality of life during the Covid-19 pandemic

**DOI:** 10.1101/2020.07.08.20148916

**Authors:** Kirsti Riiser, Sølvi Helseth, Kristin Haraldstad, Astrid Torbjørnsen, Kåre Rønn Richardsen

## Abstract

**Purpose:** First, to describe adolescents’ health information sources and knowledge, health literacy (HL), health protective measures, and health-related quality of life (HRQoL) during the initial phase of the Covid-19 pandemic in Norway. Second, to investigate the association between HL and the knowledge and behavior relevant for preventing spread of the virus. Third, to explore variables associated with HRQoL in a pandemic environment.

**Methods:** This cross-sectional study includes survey data from 2,205 Norwegian adolescents 16–19 years of age. The participants reported on their health information sources, HL, handwashing knowledge and behavior, number of social interactions, and HRQoL. Associations between study variables and specified outcomes were explored using multiple linear and logistic regression analyses.

**Results:** Television (TV) and family were indicated to be the main sources for pandemic-related health information. Handwashing, physical distancing, and limiting the number of social contacts were the most frequently reported measures. HL and handwashing knowledge and HL and handwashing behavior were significantly associated. For each unit increase on the HL scale, the participants were 5% more likely to socialize less with friends in comparison to normal. The mean HRQoL was very poor compared to European norms. Being quarantined or isolated and having confirmed or suspected Covid-19 were significantly negatively associated with HRQoL, but seeing less friends than normal was not associated. HL was significantly positively associated with HRQoL, albeit of minor clinical importance.

**Conclusion:** Adolescents follow the health authorities’ guidelines and appear highly literate. However, high fidelity requires great sacrifice because the required measures seem to collide with certain aspects that are important for the adolescents’ HRQoL.

**Implications and contribution:** This study is among the first to investigate health information sources and knowledge, health literacy, protective measures, and health-related quality of life (HRQoL) among adolescents during the Covid-19 pandemic. It identifies that the participating adolescents have high health literacy and knowledge about preventing the spread of the virus but that, at the same time, their HRQoL is poor.

## Introduction

During the first half of 2020, the coronavirus has spread around the world and the World Health Organization (WHO) declared this situation as a pandemic (1). Effective responses to the challenge posed by a virus that is as contagious as corona entail massive public involvement through basic protective measures, such as practicing hand hygiene and social distancing (2). In order to be successful, these measures require that adequate information and advice be provided so that individuals understand what they need to do, how to follow instructions and guidance, and how to ultimately make effective decisions related to their own health and the health of others. Health communication that is meant to educate people about coronavirus and how to avoid the disease or its spread has become widely available.

Adolescents constitute one important target group because they are increasingly independent and responsible for their own health behaviors. Even though there is a lower likelihood of younger people becoming ill—and despite the frequent lack of symptoms in those who do—they can nevertheless spread the virus (2). Given the critical role of person-to-person transmission in the spread of coronavirus and the fact that adolescents socialize in close peer groups, behavioral compliance may be of particular importance for this group.

Health literacy (HL) can have an impact on the effective use of health knowledge. A much applied definition by Nutbeam (3) states that HL consists of functional, interactive, and critical domains. The functional domain refers to basic skills for reading and writing health information, the interactive domain represents advanced skills that allow individuals to extract and derive meaning from health information while the critical domain refers to advanced skills used to critically evaluate health information and take control over health determinants (3). Although HL among young people has gained increasing attention, the topic is under-researched in comparison to research conducted on the adult population (4). To our knowledge, there are no existing studies on the role of adolescent HL in prevention of infectious diseases. During a pandemic, it is imperative that information is not only timely and accurate, but that it is also tailored to different populations. In the age of digital technology and easy access to information, adolescents use social media frequently for many reasons—including to search for information about health-related questions (5, 6).

The Covid-19 pandemic has resulted in lockdowns worldwide, and schools and leisure activities have been closed. In this environment, health-related quality of life (HRQoL) in adolescents may be negatively influenced by the rapid implementation of social distancing measures that are necessary for preventing the spread of the pandemic. HRQoL refers to how individuals subjectively assess their own well-being within several dimensions of life, including physical, psychological, and social functions (7). When institutions are closed, adolescents’ feelings of loneliness and isolation may increase. Moreover, a virus outbreak can create uncertainty and anxiety for all, including adolescents. This may very well impact the well-being of youth. However, thus far, no studies have investigated correlates of health-related quality of life of adolescents during a pandemic outbreak. Previous research has found that adolescents with low HL experience poor HRQoL (8); however, this relationship has not previously been studied in a pandemic environment.

Consequently, the first aim of this study is to describe adolescents’ health information sources and knowledge, their health literacy, health protective measures, and HRQoL during the Covid-19 pandemic. Its second aim is to investigate the association between health literacy and knowledge and the behavior relevant for preventing the spread of the virus; while its third aim is to identify variables associated with HRQoL in a pandemic environment.

## Methods

### Design and sampling

This cross-sectional study included Norwegian adolescents 16–19 years of age. The participants were recruited using snowball sampling. A link to an electronic survey was shared through the researchers’ network and social media platforms (Facebook and Twitter), via email to employees at the researcher’s faculties, via email to headmasters of junior high schools in municipalities across two counties, and by an influencer (Snapchat) who is also a public health nurse. The request was for recipients to answer the survey if they were in the eligible age group and/or to forward the link to potential participants in their network. Access to the survey began 3.5 weeks into the Norwegian lockdown and remained open for the subsequent two weeks (weeks 15 and 16 of 2020).

### Outcome measures

The following sociodemographic variables were collected: age, gender, urban or rural residency, parental education level, and whether the parents were born in Norway or in another country. Response categories for the item “Are you, or have you been, infected with the coronavirus?” were: “no,” “yes,” and “I suspect I am/have been.” We collapsed the two latter response categories in the analyses. Response categories for the item “How do you live currently?” were: “I am quarantined or isolated,” “I am home but not quarantined or isolated,” or “other.” The two latter categories were collapsed in the analyses.

Participants were asked to indicate the sources from which they receive information on pandemic-related protective health measures—TV, radio, newspapers, podcasts, YouTube, Snapchat, TikTok, Instagram, Facebook, other media, family, friends, school. Their existing knowledge about protective measures was collected by asking respondents to list all measures they recall (open text).

HL was measured by the Health Literacy in School-Aged Children (HLSAC) questionnaire, which is a brief scale recommended for use in large-scale studies (9, 10). HLSAC includes two items from each of the following five components: theoretical knowledge, practical knowledge, critical thinking, self-awareness, and citizenship (10). Five of the items were informed by the extensive and much applied Health Literacy Questionnaire (11). In the current study, we asked the respondents to answer the questions in light of the current pandemic situation. The HLSAC scale was originally validated for adolescents in seventh and ninth grades but it has also been applied in studies on older adolescents (10, 12, 13). The scale was not previously properly validated in Norwegian; hence, we conducted exploratory factor analysis, which identified a dominant first factor with eigenvalue = 3.88. Cronbach’s alpha for the scale was 0.86, indicating good internal consistency.

HRQoL was assessed using KIDSCREEN-10, which is a generic 10-item unidimensional instrument focusing on the functional, mental, and social aspects of well-being in children and adolescents 8–18 years of age. It is frequently applied worldwide and is particularly suited for larger epidemiological studies (14). The instrument consists of the following items starting with “thinking of last week, have you… 1) felt fit and well, 2) felt full of energy, 3) felt sad, 4) felt lonely, 5) had enough time for yourself, 6) been able to do the things that you want in your free time, 7) parent(s) treated you fairly, 8) had fun with your friends, 9) got on well at school, 10) been able to pay attention at school?” There are five response categories for each item, ranging from never to always or from not at all to extremely. The answers are recoded so that higher values always indicate better well-being. The measure is found to be valid and reliable (15, 16). In this study, Cronbach’s alpha was 0.79, showing an acceptable internal consistency for the questionnaire.

We were unable to find an instrument to measure hand hygiene in adolescent. As a result, we constructed an eight-item hand hygiene questionnaire on handwashing knowledge and behavior. Each item was measured on a five-level ordinal scale (“completely disagree”, “disagree”, “neither disagree or agree”, “agree” and “completely agree”). Exploratory factor analysis with varimax rotation identified that three items loaded on one factor reflecting hand hygiene knowledge (factor loadings 0.60–0.67) and three items loaded on one factor reflecting hand hygiene behavior (factor loadings 0.58–0.72). The remaining two items (“I engage in handwashing without thinking” and “I strive to follow handwashing advice”) did not load on any specific factor, had factor loadings of <0.5, thus, were not included in further analyses. Handwashing knowledge (“I know when to wash my hands,” “I know how to wash my hands properly,” and “I find handwashing advice easy to understand”) and handwashing behavior (“I wash my hands before socializing,” “I wash my hands after socializing,” and “I remind others to wash their hands”) were operationalized as the sum score of the three loading items. The two scales showed an acceptable internal consistency with a Cronbach’s alpha of 0.75 (handwashing knowledge) and 0.76 (handwashing behavior).

Finally, the respondents were asked to report how many different friends they met with during the last week, and social distancing was measured by asking whether they spend time with more or fewer friends than usual. The social distancing response categories were: “fewer friends,” “more friends,” and “equal number of friends.” The two latter categories were collapsed in the analyses.

### Ethical considerations

Data was collected through an anonymous web survey, “Nettskjema” (17). The study was presented to the Norwegian Regional Committee for Medical and Health Research Ethics. The Committee concluded that the study is not covered by the Health Research Act because the project does not make use of or bring forth new personal health information or sensitive data. In order to ensure full anonymity, we used broad categories for sociodemographic variables.

### Data analysis

Continuous data are presented as means with standard deviation (SD) and as medians with interquartile range (IQR), as appropriate. Categorical data are presented as counts (n) and relative frequencies (%). Crude differences between demographic groups in terms of health literacy, handwashing knowledge, handwashing behavior, and social distancing were analyzed using the Kruskal–Wallis test with post-hoc Mann–Whitney U tests and the Chi square test. Associations among the study variables and specified outcomes were explored using multiple linear regression and multiple logistic regression. Due to a technical error, a single HLSAC item was initially unintentionally omitted from the web survey and inserted after 443 survey submissions. This resulted in a substantial number of missing values on this item (19.7%). Consequently, we performed multiple imputation by chained equations to impute the HLSAC sum score, utilizing demographic variables and other HLSAC items predictive of missing or the item value as auxiliary variables in the imputation model (18). We did not impute values for other variables with missing values except for the initially omitted item due to the limited number of missing values; the KIDSCREEN-10 sum score was missing for 4.2% and the social distancing score was missing for 2.2% of the participants. For the remaining variables, the missing values were observed in 0–0.7% of the participants. Multiple regression analyses were based on the pooled estimates from 100 imputed datasets (19) and p-values of <.05 were considered to be statistically significant. All analyses were performed using the STATA 15.0. NVivo12 (QSR International) software was used to sort the responses to the open-ended questions on knowledge of protective measures. We sorted and summarized all the responses in subcategories before merging them into broader categories according to the official national recommendations (20).

## Results

### Characteristics of the sample

In total, 2,205 adolescents responded to the survey, of which 82.5% were girls and 17.2% were boys. The mean age of respondents was 17.3 (SD 1.1). While the great majority were currently well, 0.5% reported that they had been or were currently infected by the virus and 7.4% suspected that they had been or were currently infected. Almost all the adolescents (90.8%) stayed at home with social restrictions, while 4.2% were in insolation or quarantine (Table 1).

**Table 1.** Characteristics of the sample

|  |  | N (%) |
| --- | --- | --- |
| Total sample |  | 2,205 |
| Gender | *Girls* | 1,819 (82.5) |
|  | *Boys* | 379 (17.2) |
|  | *Not answered* | 7 (0.3) |
| Age | *16 yrs.* | 690 (31.4) |
|  | *17 yrs.* | 545 (24.8) |
|  | *18 yrs.* | 535 (24.3) |
|  | *19 yrs.* | 429 (19.5) |
|  | *Not answered* | 6 (0.3) |
| Health status | *Currently well* | 2,030 (92.1) |
|  | *Is/Has been infected* | 10 (0.5) |
|  | *Suspects infection* | 164 (7.4) |
|  | *Not answered* | 1 (.05) |
| Living situation | *Home isolation/quarantine* | 92 (4.2) |
|  | *Home with regulations* | 2,002 (90.8) |
|  | *Other* | 111 (5.0) |
|  | *Not answered* | 0 |

The sample consisted of 51.1% urban residents and 48.9% rural residents. Altogether, 87.8% of respondents reported that their mother was born in Norway, while 11.6% reported that their mother was born in another country. The corresponding percentages for fathers were 88.7% and 10.5%, respectively. Relative frequencies of parental educational attainment showed that 68.1% of mothers and 58.1% of fathers had higher education, while 19.6% of mothers and 25.6% of fathers had upper secondary/tertiary education. Educational attainment level below upper secondary education was reported for 3.8% and 5.8% of mothers and fathers, respectively.

### Pandemic-related health information and knowledge

The mean number of information sources accessed was 6.1 (2.8). There was no significant difference in the number of sources between groups defined by mother’s or father’s educational level nor between groups defined by either mother’s country of birth or father’s country of birth. TV (86%) and family (81.1%) were indicated to be the main sources of pandemic-related health information, while 58.6% also reported reading newspapers for information. Snapchat (59.8%) and Facebook (51.1%) were the most frequently used social media platforms. Altogether 1,972 (89.4%) of the 2,205 participants responded to the open-ended question on knowledge about protective measures, providing an average of 4.09 suggestions each. Handwashing, physical distancing, and limiting the number of social contacts were the most frequently reported known measures (Figure 1).

**Figure 1.**
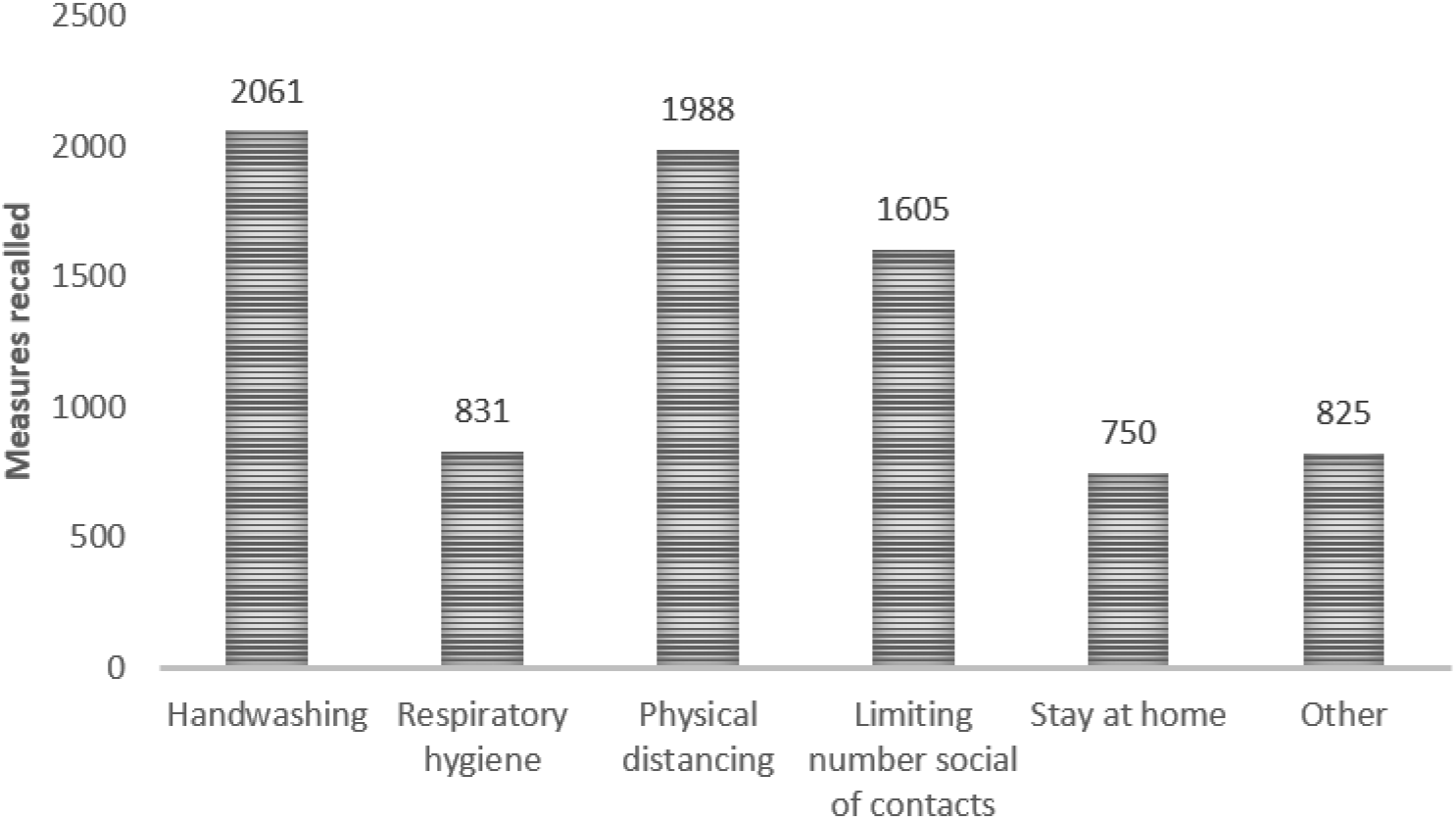
Count of health protective measures recalled^§^. ^§^“Handwashing” includes the washing of hands with soap and/or antibacterial agents; “Physical distancing” includes keeping distance and avoiding handshaking, hugging, etc.; “Respiratory hygiene” refers to covering mouth and nose with bent elbow or tissue when coughing or sneezing”; “Other” includes, e.g., the use of gloves and not touching the face. The number of measures exceeds the number of participants because some mentioned different measures that were collapsed into the same category, e.g. handwashing with soap *and* antibacterial agents.

### Health literacy and protective health measures

The overall sample scores and unadjusted group differences in HL, handwashing knowledge and behavior, and social distancing are reported in Table 2. The total sample mean score for HL was 35.2. The mean score for handwashing knowledge and behavior was 14.1 (1.6) and 11.9 (2.6), respectively. More specifically, with regards to handwashing behavior, 54.2 % of the girls and 46.9% of the boys reported reminding others to wash their hands (agree/completely agree), 72.3 % of the girls and 66.7% of the boys agreed or completely agreed that they washed their hands before socializing, and, 88.9% of the girls and 80.8 % of the boys agreed or completely agreed that they washed their hands after socializing. The gender differences were statistically significant (p<0.05). Overall, 86.4% of the sample reported currently spending time with fewer friends than they normally would. With respect to the reported number of friends seen during one week after the lockdown was implemented, the sample median value (IQR) was 2 (1–3). HL group differences (crude) were observed across age cohorts (*p* < 0.01). A significantly larger proportion of urban residents reported social distancing (88.3%) in comparison to rural residents (84.4%). Adolescents who did not know their mother’s educational level had a significantly lower HL score than those who reported any other educational level (*p* < 0.05). This finding was replicated with respect to the father’s educational level (*p* <0.01). Social distancing was reported by a smaller proportion of those who responded that their mother’s or father’s education was “below secondary” (*p* < 0.01) or “don’t know/not relevant” (*p* < 0.01) in comparison to the respondents who indicated “university/college” (Table 2).

**Table 2.** Health literacy, handwashing knowledge and behavior, and social distancing across subgroups

|  |  | Health literacy <sup>a</sup> |  | Handwashing knowledge <sup>b</sup> |  | Handwashing behavior <sup>c</sup> |  | Social distancing |  |
| --- | --- | --- | --- | --- | --- | --- | --- | --- | --- |
|  |  | Mean (SD) | *P* <sup>§</sup> | Mean (SD) | *P* <sup>§</sup> | Mean (SD) | *P* <sup>§</sup> | % | *P* <sup>§§</sup> |
| Total sample |  | 35.2 (4.0) |  | 14.1 (1.6) |  | 11.9 (2.6) |  | 86.4 |  |
| Gender | *Girls* | 35.3 (3.8) | 0.11 | 14.1 (1.5) | <0.01 | 12.0 (2.4) | <0.01 | 87.3 | 0.02 |
|  | *Boys* | 34.7 (4.6) |  | 13.7 (1.7) |  | 11.3 (2.9) |  | 82.5 |  |
| Age | 16 | 34.7 (3.9) | <0.01 | 13.9 (1.7) | <0.01 | 11.8 (2.6) | <0.01 | 87.0 | 0.16 |
|  | 17 | 35.1 (3.1) |  | 14.1 (1.4) |  | 11.7 (2.5) |  | 88.7 |  |
|  | 18 | 35.6 (3.9) |  | 14.0 (1.7) |  | 12.0 (2.6) |  | 85.4 |  |
|  | 19 | 35.9 (4.0) |  | 13.4 (1.2) |  | 12.2 (2.5) |  | 83.9 |  |
| Corona infection | *No* | 34.5 (4.9) | 0.20 | 14.1 (1.6) | 0.69 | 11.9 (2.6) | 0.63 | 86.5 | 0.49 |
|  | *Confirmed/Suspected* | 35.3 (3.9) |  | 14.0 (1.6) |  | 11.9 (2.6) |  | 84.6 |  |
| Living situation | *Isolation/Quarantine* | 35.9 (3.5) | 0.14 | 14.2 (1.3) | 0.30 | 12.3 (2.6) | 0.09 | 85.7 | 0.85 |
|  | *Not* | 35.2 (3.9) |  | 14.1 (1.6) |  | 11.9 (2.6) |  | 86.4 |  |
|  | *Isolation/Quarantine* |  |  |  |  |  |  |  |  |
| Residency | *Urban* | 35.5 (3.9) | <0.01 | 14.1 (1.5) | 0.93 | 12.0 (2.5) | 0.03 | 88.3 | <0.01 |
|  | *Rural* | 35.0 (4.0) |  | 14.0 (1.7) |  | 11.8 (2.6) |  | 84.4 |  |
| Mother’s education | *Below secondary* | 35.0 (4.7) | <0.01 | 14.0 (1.7) | 0.20 | 11.7 (2.8) | <0.01 | 76.8 | <0.01 |
|  | *Higher secondary* | 35.0 (4.2) |  | 14.2 (1.6) |  | 12.2 (2.6) |  | 86.1 |  |
|  | *University/College* | 35.5 (3.7) |  | 14.1 (1.5) |  | 11.8 (2.6) |  | 87.8 |  |
|  | *Don’t know/Not relevant* | 33.8 (4.7) |  | 14.0 (1.7) |  | 11.8 (2.7) |  | 86.4 |  |
| Father’s education | *Below secondary* | 35.3 (4.7) | <0.01 | 14.1 (1.7) | 0.07 | 12.0 (2.8) | <0.01 | 77.3 | <0.01 |
|  | *Higher secondary* | 35.2 (3.9) |  | 14.2 (1.6) |  | 12.1 (2.5) |  | 86.1 |  |
|  | *University/College* | 35.5 (3.7) |  | 14.1 (1.5) |  | 11.8 (2.6) |  | 88.2 |  |
|  | *Don't know/Not relevant* | 33.7 (4.4) |  | 13.9 (1.7) |  | 11.8 (2.6) |  | 81.8 |  |
| Mother's country of birth | Norway | 35.2 (3.9) | 0.58 | 14.1 (1.5) | 0.14 | 11.9 (2.6) | 0.90 | 86.4 | 0.99 |
|  | Other country | 35.2 (4.5) |  | 13.9 (1.7) |  | 11.9 (2.7) |  | 86.4 |  |
| Father's country of birth | Norway | 35.2 (3.8) | 0.41 | 14.1 (1.6) | 0.27 | 11.9 (2.6) | 0.72 | 86.6 | 0.48 |
|  | Other country | 35.2 (4.6) |  | 14.0 (1.5) |  | 11.9 (2.6) |  | 85.0 |  |

<sup>§</sup> Kruskal–Wallis mean ranks test (>2 group comparison) or Mann–Whitney U test (2 group comparison) <sup>§§</sup> $\chi^2$ test.
<sup>a</sup> Sum score obtained from the Health Literacy in School-Aged children (HLSAC) questionnaire [min–max: 10–40]. Higher score reflects higher level of health literacy. | <sup>b</sup> Sum score of three items [min–max: 3–15]. Higher score reflects higher level of knowledge. | <sup>c</sup> Sum score of three items [min–max: 3–15]. Higher score reflects higher level of protective behavior.

General HL was statistically associated with handwashing knowledge (0.14; 95% CI [.15:.21]) and handwashing behavior (0.18; 95% CI [.15:.21]). Moreover, for each additional unit on the HLSAC scale, the participants were 5% more likely to socialize less with friends in comparison to normal (OR = 1.05; 95% CI [1.01:1.08]) (see Table 3). Sensitivity analysis performed with complete data confirmed our findings from the imputed data set.

**Table 3:** Health literacy association with hand hygiene knowledge and hygiene behavior and social distancing

|  | **Model 1<sup>a</sup>**<br>Association between health literacy and hand hygiene knowledge (n = 2,022) |  | **Model 2<sup>a</sup>**<br>Association between health literacy and hand hygiene behavior (n = 2,024) |  | **Model 3<sup>b</sup>**<br>Association between health literacy and social distancing (n = 1,999) |  |
| --- | --- | --- | --- | --- | --- | --- |
| | $\beta$ | 95% CI | $\beta$ | 95% CI | Odds ratio | 95% CI |
| Health literacy <sup>c</sup> | .14*** | .12 - .15 | .18*** | .15 - .21 | 1.05** | 1.01 - 1.08 |

<sup>a</sup> Multiple linear regression analysis with control for gender, maternal education, paternal education, mother's country of birth, father's country of birth, and residence type (urban vs rural). Pooled estimates from 100 imputed datasets.
<sup>b</sup> Multiple binary logistic regression analysis with control for gender, maternal education, paternal education, mother's country of birth, father's country of birth, and residence type (urban vs rural). Pooled estimates from 100 imputed datasets.
<sup>c</sup> Sum score obtained from the Health Literacy in School-Aged Children (HLSAC) questionnaire [min–max: 10–40]. Higher score reflects higher level of health literacy.
\*\* $p < .01$ . \*\*\* $p < .001$

### Variables associated with health-related quality of life

The sample mean value (SD) for KIDSCREEN-10 was 55.3 (17.0). The results of the analysis of variables associated with HRQoL are presented in Table 4. The analysis showed a statistically significant association between HRQoL and HL 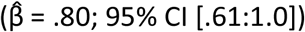. In addition, the analysis confirmed that being a boy was statistically positively associated with HRQoL 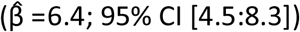, and having confirmed or suspected Covid-19 was statistically negatively associated with HRQoL 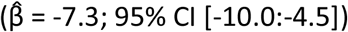. While being isolated or in quarantine was associated with lower HRQoL 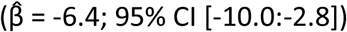, there was no difference between those who reported they spent time with fewer friends the last week and those who spent time with an equal number, or more friends. In contrast to having a father with higher education, having a father with below secondary education 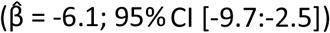or higher secondary education level 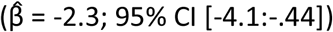 were both negatively associated with HRQoL.

**Table 4:** Variables associated with health-related quality of life (HRQoL)^a^ (n=1,928)

| | $\beta$ | 95% CI |
| --- | --- | --- |
| Health literacy <sup>b</sup> | .80*** | .61 - 1.0 |
| Boys (ref: *Girls*) | 6.4*** | 4.5 - 8.3 |
| Age (ref: 16 yrs.) |  |  |
| 17 yrs. | -1.6 | -3.5 - .38 |
| 18 yrs. | -1.2 | -3.2 - .72 |
| 19 yrs. | -1.6 | -3.7 - .54 |
| Quarantined/Isolated (ref: *Not quarantined/Isolated*) | -6.4*** | -10.0 - -2.8 |
| Social distancing (ref: Spent time with equal number or more) |  |  |
| *Spent time with fewer friends last week* | .28 | -1.9 - 2.5 |
| Confirmed/Suspected Covid-19 (ref: *Not infected*) | -7.3*** | -10.0 - -4.5 |
| Father's education (ref: *Higher education*) |  |  |
| *Below secondary education* | -6.1** | -9.7 - -2.5 |
| *Secondary education* | -2.3* | -4.1 - -.44 |
| *Don't know/Not relevant* | -3.0 | -6.0 - .00 |
| Mother's education (ref: *Higher education*) |  |  |
| *Below secondary education* | -1.7 | -5.7 - 2.2 |
| *Secondary education* | -.09 | -2.10 - 1.9 |
| *Don't know/Not relevant* | -3.6* | -7.0 - -.20 |
| Rural residence (ref: *Urban residence*) | 1.6* | -.07 - 3.0 |
| Father's birth country (ref: *Norway*) |  |  |
| *Other country* | -.50 | -2.1 - 3.1 |
| Mother's birth country (ref: *Norway*) |  |  |
| *Other country* | -.67 | -3.2 - 1.8 |

<sup>a</sup> KIDSCREEN-10 [min-max: 0-100]. Higher score reflects better health-related quality of life.
<sup>b</sup> Health Literacy in School-Aged Children [sum score range: 10-40]. Higher score reflects higher level of health literacy.
\* $p < .05$ . \*\* $p < .01$ . \*\*\* $p < .001$

## Discussion

In this study, we intended to obtain deeper insight about adolescents’ sources of health information and their protective health knowledge and behavior in specific relation to the Covid-19 pandemic and how these variables are associated with their health literacy. Moreover, we aimed to explore the association between various variables, including those characterizing the pandemic environment and the health-related quality of life of adolescents.

The findings show that the participating adolescents use several different sources to stay informed and that three to four weeks into the lockdown they were well educated about the protective health measures necessary to prevent the virus from spreading. Interestingly, to remain informed, they seemed to utilize their family and traditional media as information sources more than social media platforms. The participants were conscious of the importance of handwashing and the need to limit social interactions by seeing less friends than usual. A large majority reported complying with the guideline on protective measures, both with respect to handwashing and social distancing. However, the proportion of respondents reporting compliance was higher among girls than among boys. Their HL was high and statistically significantly associated with handwashing knowledge and behavior as well as with the application of social restrictions.

The results indicate that, in the initial phases of the pandemic, health authorities succeeded in providing information that is easy for young people to understand. The participating adolescents seem to be generally well informed about protective behavior advice, with handwashing, physical distancing, and social contact limiting being the most frequently recalled recommendations. The two latter measures have the greatest impact on adolescents’ lives. In comparison to handwashing, which requires establishing a habit, the social contact limitation restrains adolescents from an activity that is completely natural, enjoyable, and even essential for them. These sudden restrictions like being quarantined seem to heavily impede the well-being of young people. The adolescents in the present study scored notably lower on HRQoL (55.3) in comparison to the sample in a previous Norwegian reference study, whose mean KIDSCREEN-10 score was 64.59 (21). The difference can be interpreted as being clinically important (22). In comparison to European norm data for adolescents, the present sample mean score was between the 12.7 and the 16.1 percentile (14). Collective efforts and being part of something important like fighting a virus, may increase a feeling of connectedness, however, complying with advice also comes with a price. Taking the current circumstances into consideration, it may not come as a surprise that the adolescents scored lower on social life, leisure time, and autonomy items. It can also be argued that this is of minor concern because their HRQoL will most likely increase as soon as restrictions are lifted. However, since health authorities around the world signal that precautions will need to continue in the foreseeable future, the results are worrying. Even though the majority of adolescents succeed in dealing with temporary restrictions, there are reasons to be particularly attentive to vulnerable groups, whose HRQoL is already poor— e.g., adolescents with mental health problems (23), chronic diseases (24), or adolescents in families with low socioeconomic status (24, 25). The explorative nature of this study allowed us to investigate possible correlates of the adolescents’ HRQoL under the present circumstances. The results are very much in line with previous research, which shows that HRQoL is associated with gender (boys report higher HRQoL compared to girls) and parental level of education (15, 26). Unsurprisingly, being infected with the virus or being isolated or quarantined was negatively associated with HRQoL. Spending time with friends is found to be essential for adolescents’ quality of life (27). Thus it may seem contradictory that there was no difference in HRQoL between those who reported that they spent time with fewer friends and those who spent time with an equal number, or more friends. An explanation for this may be that although the majority socialized less, they kept in touch with a few significant friends. Another reasonable explanation could be that the ones spending time with an equal number of friends, have few friends and lower HRQoL also under normal circumstances.

Interestingly, HL was statistically associated with HRQoL, but the association was small and must be seen more as a tendency. A probable explanation for this is that the HLSAC instrument caused a ceiling effect. According to cut points proposed, the sample and subgroup HL mean scores were medium and close to high (10, 28). This could also be caused by the age range of the present sample. However, the adolescents in our study scored higher in comparison to Norwegian secondary school students in a previous study (12). The sample proportion of participants with parents who have higher education levels was larger in comparison to the general Norwegian population (34,1%), which may also partly explain the high HL scores (29). However, there were only minor differences in HL scores between participants with high versus low parental education. It is likely that the overall high HL scores reflect that the adolescents were asked to respond to the HL questions in light of the current pandemic situation. Furthermore, pandemic-related information has constantly been communicated through conventional and social media since the outbreak in Norway. A change in collective behavior, consciousness and alertness is most probable a result of a massive health information uptake.

### Limitations

A pandemic such as the current one has not been experienced before. Hence, there was little previous research to lean on in the planning and execution of the present study. Time was an important factor and it was not possible to pilot the method for data collection. The recruitment strategy resulted in a sample that is less representative of boys and adolescents from families with lower educational levels thus affecting the generalizability to Norwegian youth in general with respect to prevalence of health protective behavior and levels of health literacy and HRQoL. To reduce the response burden, we had to make several pragmatic choices with regard to applied instruments. More comprehensive instruments with subdomains would have given more information on HL and HRQoL aspects not covered here. This study does not include data on variables that more specifically can explain which restrictions (e.g. closing of schools or leisure activities) that affects dimensions of HRQoL. Furthermore, the lack of validated instruments on specific protective health measures (hand hygiene) forced us to rapidly design an instrument that is not yet being validated. Missing values on one HLSAC item were observed, especially among boys. The technical error most likely did not lead to any biases; however, multiple imputation was performed in order to strengthen the plausibility of the missing at random assumption.

## Conclusion

The present study is among the first to describe health information sources and knowledge, health literacy, health protective measures, and HRQoL among adolescents during the Covid-19 pandemic. The participating adolescents appeared to be highly literate and motivated to follow the health authorities’ guidelines. However, high fidelity requires great sacrifice because the necessary preventive measures collide with aspects that are important for the quality of life of adolescents.

## Data Availability

Data will be made available after acceptance and submission

## Acknowledgements

The authors are grateful for statistical advice from Dr Milada Cvancarova Småstuen at Oslo Metropolitan University, Faculty of Health Sciences.

